# Psychological, endocrine and polygenic predictors of emotional well-being during the COVID-19 pandemic in a longitudinal birth cohort

**DOI:** 10.1101/2022.02.03.22270311

**Authors:** Thao Nguyen, Lea Zillich, Metin Cetin, Alisha S.M. Hall, Jerome C. Foo, Lea Sirignano, Josef Frank, Tabea S. Send, Maria Gilles, Marcella Rietschel, Michael Deuschle, Stephanie H. Witt, Fabian Streit

**Affiliations:** Department of Psychiatry and Psychotherapy, Central Institute of Mental Health, Medical Faculty Mannheim, University of Heidelberg, J5, 68159 Mannheim, Germany; Department of Genetic Epidemiology in Psychiatry, Central Institute of Mental Health, Medical Faculty Mannheim, University of Heidelberg, J5, 68159 Mannheim, Germany

**Keywords:** children, COVID-19, emotional well-being, HPA axis, polygenic risk score, prenatal stress

## Abstract

The COVID-19 pandemic severely affected the lives of families, and the well-being of children and their parent. Prenatal stress, dysregulation of the hypothalamic-pituitary-adrenal (HPA) axis, and genetic factors might influence individuals’ well-being in the presence of a major stressor such as the COVID-19 pandemic.

The present work is part of an ongoing birth cohort study and aims to investigate maternal perceived stress, early childhood HPA axis activity, and polygenic risk scores (PRSs) as predictors of emotional well-being during the COVID-19 pandemic. All participants are part of the ongoing birth cohort study POSEIDON. Emotional well-being of children (*n*=263) and mothers (*n*=241) was assessed during the COVID-19 pandemic using the CRISIS questionnaire in two waves between June 2020 and February 2021. Associations of well-being with previously assessed maternal perceived stress, children’s salivary and morning urine cortisol at 45 months, PRSs for depression, schizophrenia, loneliness were investigated.

A positive association between the children’s and mothers’ emotional well-being was found. Lower emotional well-being was observed in both children and mothers during the pandemic compared to before. Children’s emotional well-being improved over the course of the pandemic. Prenatally assessed maternal perceived stress was associated with a decrease in children’s but not in the mothers’ well-being. Cortisol measures and PRSs were not significantly associated with emotional wellbeing.

The present study confirms that emotional well-being of children and mothers are linked, and were negatively affected by the COVID-19 pandemic, with differences in development over time.

## 1. Introduction

Stress is a major risk factor for somatic and mental health problems (Agorastos and Chrousos, 2021). The COVID-19 pandemic represents a major stressor. Findings on mental health during the COVID-19 pandemic are heterogeneous, demonstrating both increased as well as decreased mental distress over time (Bussières et al., 2021; Robinson et al., 2022). A wide range of factors could influence how children’s mental health developed under the influence of the life changes associated with the COVID-19 pandemic. These include parental well-being and stress perception, biological regulators of the stress response such as the functioning of the hypothalamic-pituitary-adrenal (HPA) axis, and genetic risk factors.

Children’s psychosocial well-being is closely related to that of their parents (Cobham et al., 2016). Parents who reported higher levels of depression and anxiety or stress during the COVID-19 pandemic also reported higher stress levels (Russell et al., 2020) and behavioral and emotional problems in their children (Cusinato et al., 2020; Spinelli et al., 2020). In addition from social influences, the regulation of the HPA axis has been identified as a potential mediator of the impact of stressful experiences. Early-life stress is linked to HPA axis dysregulation (O’Connor et al., 2021) and this effect can last into adulthood (Van den Bergh et al., 2020). Early-life stress, such as prenatal stress has been associated with impairments in development and health of children, which is partially mediated by early programming of HPA axis functioning (Van den Bergh et al., 2020). In line with this, associations between HPA axis regulation and pandemic-related stressors have been shown in children and adults (Ahrens et al., 2022; Haucke et al., 2022; Jopling et al., 2021; Perry et al., 2022).

Genetic factors play an important role in stress regulation (Kudielka et al., 2009) and mental disorders (Levinson, 2006). Genome-wide association studies (GWAS) identify common genetic variants associated with psychiatric disorders or particular traits. Based on these results, polygenic risk scores (PRSs) can be calculated in independent target samples, reflecting the polygenic risk burden of each individual (Wray et al., 2014). Recent studies demonstrated associations between PRS for depression and mental health in adults (McIntosh et al., 2019), adolescents and children (Kwong et al., 2021), including during the COVID-19 pandemic (Ahrens et al., 2022).

The present study aims to investigate emotional well-being during the COVID-19 pandemic and its association with maternal perceived stress, early childhood HPA axis activity and mental health-related PRSs. We hypothesize that:

(H1) The COVID-19 pandemic has negative short-term (H1a) and long-term (H1b) effects on the emotional well-being of children and mothers;

(H2) The mother’s emotional well-being is positively associated with the child’s emotional well-being;

(H3) Higher maternal perceived stress during pregnancy (3b) and at child’s age of 45 months (3c) is associated with a stronger decrease in the child’s (3a) and mother’s emotional well-being during the pandemic;

(H4) The child’s HPA axis regulation at 45 months of age is associated with the change in the child’s emotional well-being during the pandemic;

(H5) Higher PRSs for depression, schizophrenia and loneliness are associated with a stronger decrease in the child’s emotional well-being during the pandemic.

## 2. Materials and Methods

### 2.1 Sample

This study is part of the longitudinal birth cohort study “POSEIDON” (Pre-, Peri-, and Postnatal Stress: Epigenetic Impact on Depression) examining prenatal stress, health and development of children. At three obstetric clinics in the Rhine-Neckar Region of Germany, 410 pregnant women about 4-8 weeks prior to delivery were recruited for the first study wave (T1) from October 2010 to March 2013. Five waves have been conducted so far: during the third trimester of pregnancy (T1), within a few days after childbirth (T2), six months postpartum (T3), 45 months postpartum (T4) and during the COVID-19 pandemic, when children were 7-10 years old (T5). At T4, dropouts were replaced by 101 children and their parents. Details on the recruiting process, inclusion criteria, and sample characteristics have been described previously (Send et al., 2019a; Send et al., 2017).

The present work uses data from study waves T1, T4, and T5. In Germany, strict restrictions were enacted in March 2020 at the beginning of the pandemic. Restrictions were slowly eased starting mid-April and reinstated in November 2020. The current study wave T5 is divided into two parts. Baseline T5a took place between July and October 2020. Follow-up T5b took place between November 2020 and February 2021. A total of 482 children and their parents who were part of the POSEIDON cohort in at least one study wave were invited to participate again. Of these 482 children and parents, 264 participated. The final sample included 263 children at T5a and 169 at T5b. 241 mothers completed the questionnaire at T5a and 160 at T5b were included in the analyses. Some mothers only answered the questionnaires about their child, therefore there is a mismatch between the number of included children and mothers, for details see Figure 1. An overview of number of participants and samples included in the analysis is displayed in Figure 1.

**Fig. 1:**
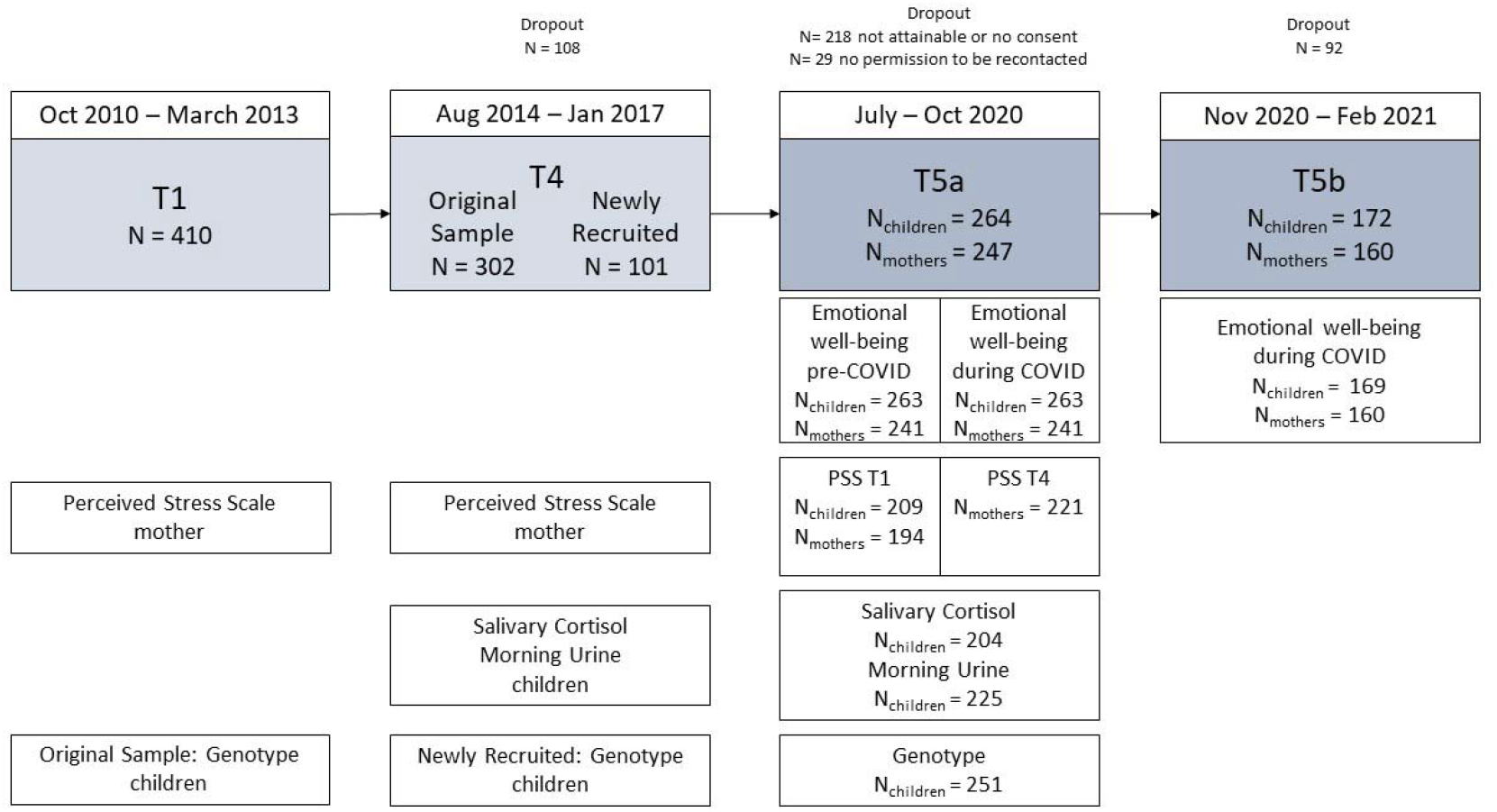
Caption: Overview of the included POSEIDON study waves: the flow diagram indicates the number of subjects participating in the POSEIDON study at the T1, T4, T5a and T5b time points, along with the number of samples that were included in the statistical analysis (N_children_ = data available for analyses of children’ well-being; N_mothers_ = data available for analyses of mothers’ well-being). Alt Text: A flow diagram shows an overview of the study waves T1, T4, T5a and T5b along with the number of subjects participating in each part and the time period in which data collection took place. The variables included in the statistical analyses and number of subsamples are displayed according to the study wave in which they were collected.

The Ethics Committee of the Medical Faculty Mannheim of the University of Heidelberg approved the study. Before participation, all families provided written informed consent.

### 2.2 Procedure

First, the children’s parents were contacted in July-August 2020 by telephone and asked whether they would be interested in participating in a short online survey. Parents who were not reachable by phone were approached by e-mail or through postal mail. If the family agreed to participate, they were provided with detailed information about the study and asked for their written informed consent. Participants who agreed to participate in the survey received an invitation by email, along with the link to the online survey and provided written consent. For participants who were either not reachable by e-mail or preferred a paper-pencil questionnaire, the questionnaires were sent by postal service. A follow-up survey was sent out between November-December 2020 to all subjects who had participated in the first online survey (T5a) and agreed to be recontacted.

The primary caregiver was asked to complete the parent/caregiver version of the CRISIS questionnaire (for measures, see 2.3.1) about the child. In addition to the parent/caregiver questionnaire, both parents were invited to answer the adult self-report version of the CRISIS. Therefore, each family received three questionnaires. The online survey was created with REDCap, a web application for building and managing online surveys (Harris et al., 2019). Study data were collected and managed using REDCap electronic data capture tools.

### 2.3 Measures

#### 2.3.1 Emotional well-being T5

Emotional well-being was assessed with the “Adult self report” and the “Parent caregiver report” versions of the emotional well-being questionnaire of the “CRISIS - The Coronavirus Health Impact Survey” (V0.2) (Merikangas et al., 2020). These questionnaires are licensed and available at crisissurvey.org. The emotional well-being scale consists of 10 items on a 5-point Likert scale, adapted from the circumplex model of affect (Larsen and Diener, 1992; Posner et al., 2005). Construct validity of the CRISIS questionnaire has been shown in other samples (Nikolaidis et al., 2021). In the first part of this study wave (T5a), children’s and parents’ emotional state three months prior to the pandemic was assessed retrospectively (referred to as T5 pre) as well as the current emotional state during the pandemic. At the second time point (T5b), the current emotional state during the pandemic was assessed. As only a relatively small proportion of fathers answered the questionnaires, the present analyses include the data from mothers and children only.

#### 2.3.2 Maternal perceived stress T1 & T4

At T1, prenatal perceived stress of the mothers was assessed with the Perceived Stress Scale (PSS) (Cohen et al., 1983). The self-report questionnaire consists of 14 items measuring the experienced level of stress during the last month (of pregnancy). Higher values regarding the sum score of all items indicate higher perceived stress. At T4 the perceived stress of the mothers was assessed again using the PSS. During the pandemic the PSS was not assessed.

#### 2.3.3 HPA axis measures T4

The nocturnal activity of the HPA axis was assessed through the cortisol concentration in the morning urine of the children at T4. Details are described elsewhere (Send et al., 2019b).

Stress reactivity was operationalized by salivary cortisol concentration before, 10, 30 and 40 minutes after finishing the stress test at T4. Briefly, the children and the researcher played a game, where the children had to attach colored magnets to a matching animal. A red stop light indicated that the time was up. The researcher switched the light remotely before the children could finish. Therefore, the children experienced failure and received negative feedback repeatedly. Full details are described elsewhere (Send et al., 2019a).

#### 2.3.4 Polygenic risk scores

For children of the original cohort, DNA was extracted from cord blood. For children newly recruited at T4, DNA was extracted from saliva using the ORAgene sampling kit (DNA Genotek, Ottawa, Ontario, Canada). The Illumina Psych Array, V1.0 for original cohort and V1.3 for newly recruited children (Illumina, Inc., San Diego, CA, USA), was used for genome-wide genotyping. Quality control and filtering was performed using PLINK 1.9 (Chang et al., 2015), according to recommendations published in Turner et al. (Turner et al., 2011). We removed participants with a mismatch between phenotypic and genotypic sex, > 0.02 missingness, or a heterozygosity rate > |0.2|. We removed SNPs with a minor allele frequency (MAF) of < 0.01, missing data > 0.02, or deviating from Hardy-Weinberg equilibrium (HWE) with a p-value < 10^−6^.

A SNP set filtered for high quality SNPs (MAF > 0.20, missingness = 0, HWE p > 0.02) and LD pruning (pairwise r^2^ < 0.1 within a 200 SNP window) was used to filter for relatedness and population structure and cryptically related (π □ > 0.20) subjects were excluded at random. Principal components were generated to control for population stratification and to remove outliers (> 6 SD on one of the first 20 principal components). After quality control, genotypic data was available for 229 subjects, who also had T5 data.

PRSice 2.1.6 (Choi and O’Reilly, 2019) was used to calculate PRS, including GWAS SNPs with an info score > 0.9, and using a p-value threshold of nominal significance (p < 0.05). Besides PRSs for depression (PRS-Depression), we calculated PRS for schizophrenia (PRS-Schizophrenia) and loneliness (PRS-Loneliness), which have previously been linked to depressive symptoms and well-being (Day et al., 2018; Okbay et al., 2016). The PRS-Depression was based on GWAS summary statistics from Howard (Howard et al., 2019), excluding German samples, the PRS-Schizophrenia, which was based on Ripke (Ripke et al., 2014) and the PRS-Loneliness was based on Day (Day et al., 2018).

### 2.4 Statistical analysis

All statistical analyses were performed in the R statistical environment version 3.5.1.

For emotional well-being, inverted items were recoded and mean scores were calculated for the three time points, with higher value corresponding to better emotional well-being (range 1 - 5). Cronbach’s alpha was calculated for the CRISIS emotional well-being scale for children and mother at each assessed time-point (*range* 0.82 – 0.88). For perceived stress, a total sum score of the PSS was calculated (Cohen et al., 1983). If only one item was missing, it was impputed with the mean of the other 13 items. Internal reliability was determined by calculating Cronbach’s alpha at T1 (0.88) and T4 (0.85).

Morning urinary cortisol concentrations were corrected for urinary creatinine concentration by dividing cortisol by creatinine concentration. The urinary cortisol concentrations corrected for creatinine were log10-transformed to reduce skewness. The salivary cortisol concentrations at the four time points were also log10-transformed to reduce skewness. The area under the curve with respect to increase (AUCi) was calculated (Pruessner et al., 2003) in relation to baseline cortisol concentration. AUCi reflects the course of cortisol concentration in response to stress induction. Outliers three standard deviations above or below the mean were winsorized to the closest value within three standard deviations.

To investigate the emotional well-being over time, repeated measures ANOVA was calculated and differences between the time points were investigated using post-hoc t-tests (H1). Pearson correlations were performed to test associations between the child’s and the mother’s emotional well-being (H2). Linear multiple regression analysis was used to investigate the relationship between perceived stress as independent variable and the change in the emotional well-being of the child (H3a) and the mother (H3bc) as dependent variable. For maternal perceived stress, the child’s sex (H3a) and age (3abc) were added as covariates because of possible influences of these variables on emotional well-being.

Investigating the relationship between the child’s AUCi, the child’s urinary cortisol level as independent variables and the change in the emotional well-being as dependent variable (H4) linear multiple regression analysis was used. For the AUCi, time of measurement was added as a covariate, for urinary cortisol the time of urine sample and whether the children wore a diaper was included because of possible influences of these variables on cortisol levels.

Linear regression models were calculated to explore an association between the PRS-Depression, PRS-Schizophrenia, PRS-Loneliness as independent variables and the change in the emotional well-being during the pandemic as the dependent variable, correcting for the first five genetic principal components, age and sex (H5).

The significance level was set to p < 0.05. Subjects were excluded pairwise from the respective analyses.

A post-hoc power analysis was calculated for the primary hypothesis, emotional well-being of children, using GPower 3.1.9.7 (Faul et al., 2009). With a total sample size of *n* = 169, an observed effect of *f* = 0.24, a mean correlation of repeated measures of *r* ∼ 0.6 and three measurements, the achieved power for this analysis was > 0.99.

## 3. Results

### 3.1 Descriptive statistics

An overview of used variables is provided in Table 1. From a total of *N* = 264 children and *N* = 247 mothers, *n* = 1 child and *n* = 6 mothers were excluded from the analyses in T5a due to missing values. From a total of *N* = 172 children and *N* = 160 mothers, *n* = 3 children were excluded from the analyses in T5b due to missing values. Mean age of the children at T5a was *M* = 7.75 (*SD* = 0.71) years and at T5b *M* = 8.02 (*SD* = 0.76) years. Mean age of the mothers at T5a was *M* = 40.73 (*SD* = 4.49) years and at T5b *M* = 40.94 (*SD* = 4.62) years. Data on morning urine cortisol was available for *n* = 226 children and data on salivary cortisol (AUCi) for *n* = 205 children. Regarding maternal perceived stress, data from *n* = 210 mothers for T1 and *n* =241 for T4 was included in the sample.

**Table 1.**
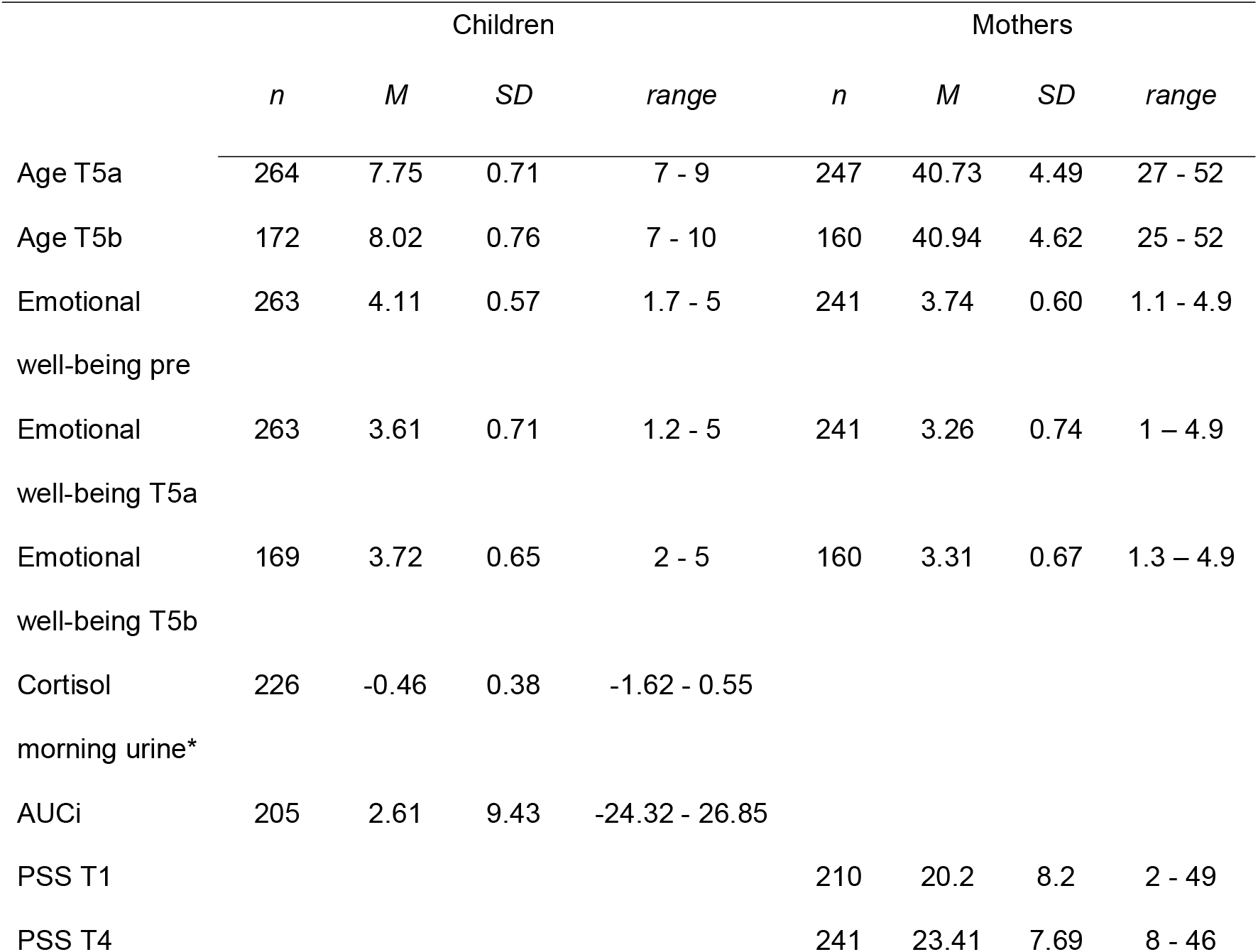
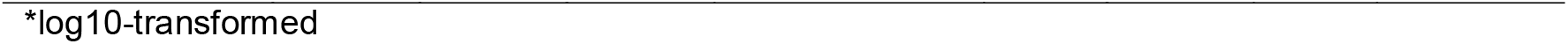
Descriptive statistics

|  | Children |  |  |  | Mothers |  |  |  |
| --- | --- | --- | --- | --- | --- | --- | --- | --- |
|  | *n* | *M* | *SD* | *range* | *n* | *M* | *SD* | *range* |
| Age T5a | 264 | 7.75 | 0.71 | 7 - 9 | 247 | 40.73 | 4.49 | 27 - 52 |
| Age T5b | 172 | 8.02 | 0.76 | 7 - 10 | 160 | 40.94 | 4.62 | 25 - 52 |
| Emotional well-being pre | 263 | 4.11 | 0.57 | 1.7 - 5 | 241 | 3.74 | 0.60 | 1.1 - 4.9 |
| Emotional well-being T5a | 263 | 3.61 | 0.71 | 1.2 - 5 | 241 | 3.26 | 0.74 | 1 - 4.9 |
| Emotional well-being T5b | 169 | 3.72 | 0.65 | 2 - 5 | 160 | 3.31 | 0.67 | 1.3 - 4.9 |
| Cortisol morning urine* | 226 | -0.46 | 0.38 | -1.62 - 0.55 |  |  |  |  |
| AUCi | 205 | 2.61 | 9.43 | -24.32 - 26.85 |  |  |  |  |
| PSS T1 |  |  |  |  | 210 | 20.2 | 8.2 | 2 - 49 |
| PSS T4 |  |  |  |  | 241 | 23.41 | 7.69 | 8 - 46 |

\*log10-transformed

### 3.2 H1: Emotional well-being in course of the pandemic

The children’s emotional well-being differed before and over the course of the pandemic, as shown by a significant ANOVA (*F*(2,336) = 76.40, *p* < 0.001), as depicted in Figure 2. A worse emotional well-being during the pandemic (T5a) compared to before the pandemic (T5 pre) was observed (*t*(168) = 12.0, *p* < 0.001). In the course of the pandemic (T5b) the emotional well-being of the children improved compared to the beginning of the pandemic (T5a) (*t*(168) = -2.77, *p* = 0.006), but is still worse compared to before the pandemic (*t*(168) = 8.88, *p* < 0.001).

**Fig. 2.**
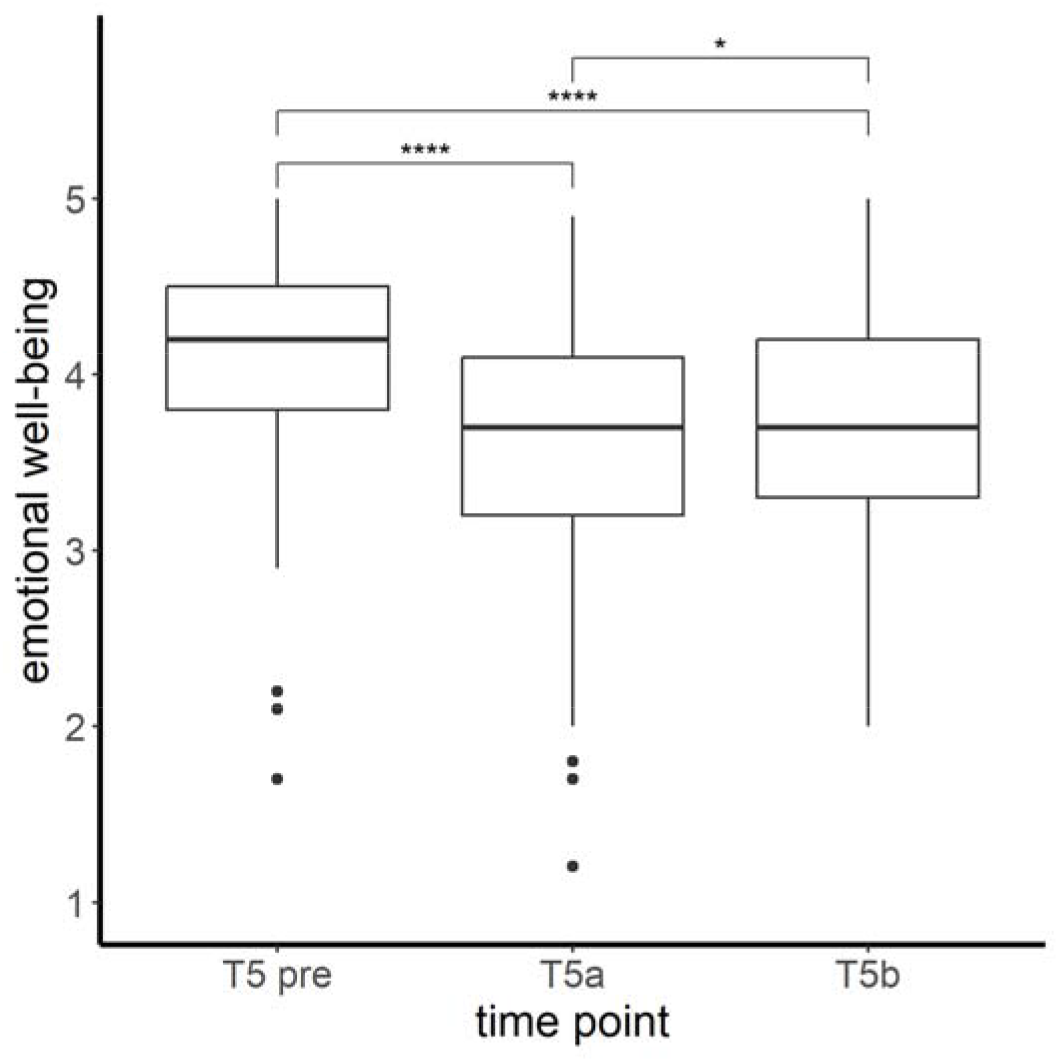
Caption: Mean levels and range of emotional well-being (CRISIS score) of the children at all three assessed time points. Significant differences in post hoc tests **** *p* < 0.001; * *p* < 0.05 Alt Text: Mean levels and range of emotional well-being (CRISIS score) of the children at all three assessed time points are shown in a boxplot. The emotional well-being was highest at the first time point and lowest at the second time point.

The same overall effect of emotional well-being was observed for mothers as the ANOVA showed significant differences in the mothers’ emotional well-being before and during the pandemic (*F*(2,314) = 67.55, *p* < 0.001), as depicted in Figure 3. A worse emotional well-being was observed comparing before the pandemic (T5a pre) to during the pandemic at T5a (*t*(157) = 9.50, *p* < 0.001) and T5b (*t*(157) = 10.4, *p* < 0.001). In the course of the pandemic (T5b), the emotional well-being of the mothers did not change significantly (*t*(157) = -0.45, *p* = 0.65).

**Fig. 3.**
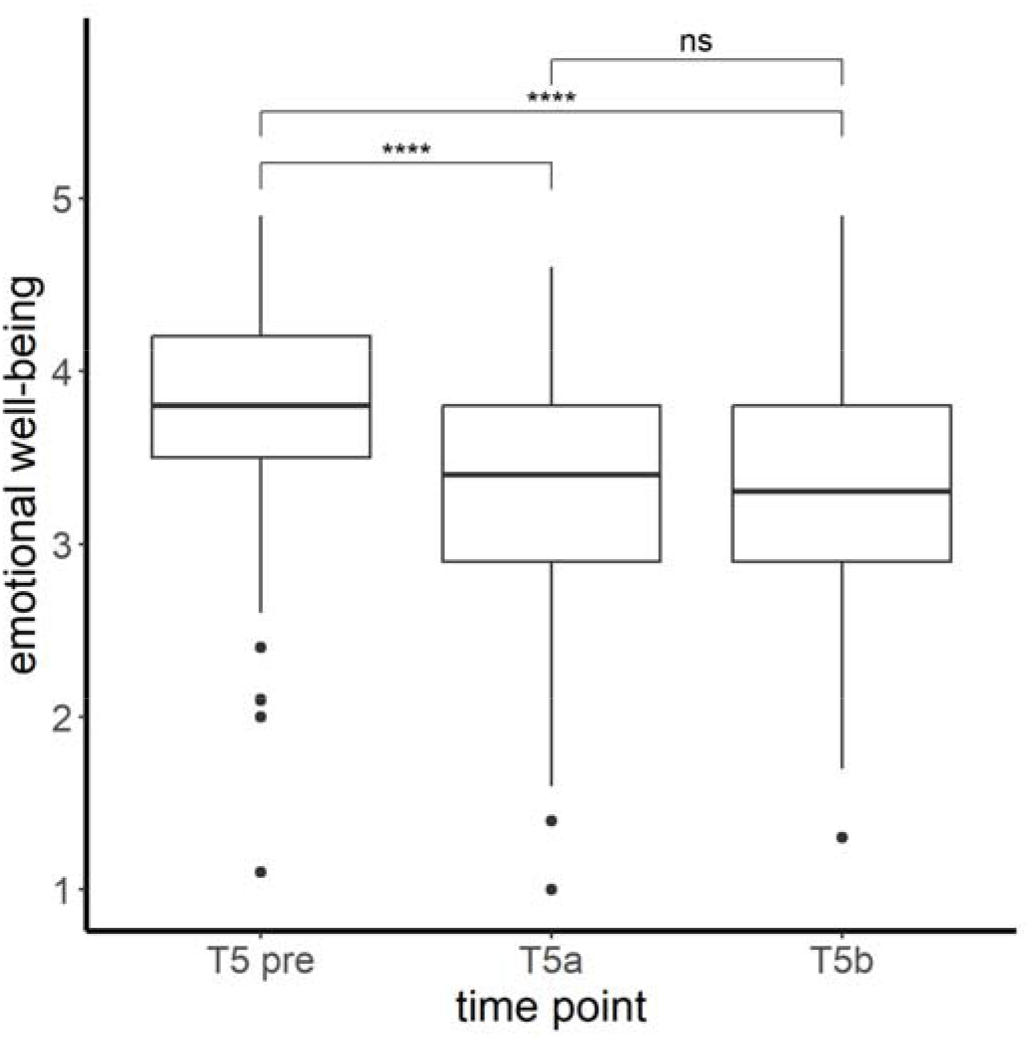
Caption: Mean levels and range of emotional well-being (CRISIS score) of the mothers at all three assessed time points. Significant differences in post hoc tests **** *p* < 0.001; ns= not significant Alt Text: Mean levels and range of emotional well-being (CRISIS score) of the mothers at all three assessed time points are shown in a boxplot. The emotional well-being was highest at the first time point and significantly lower at the second and third time point, which do not differ significantly.

### 3.3 H2: Association between the mothers’ and the child’s emotional well-being

The emotional well-being of the mother and child was positively associated at all time points, as shown by correlation coefficients of moderate size: T5 pre (*n* = 241, *r* = 0.43, *p < 0.001*), T5a (*n* = 241, *r* = 0.59, *p* < 0.001) and T5b (*n* = 156, *r* = 0.49, *p* < 0.001).

### 3.4 Association between maternal perceived stress and change in the emotional well-being

#### H3a

The model predicting the change in the child’s emotional well-being during the pandemic with prenatal perceived stress of the mother at T1, age and sex of the child explained 6% of the variance of the change in the child’s emotional well-being (*F*(3, 205) = 4.58, *p* = 0.004, *R2* = 0.063). Regression parameters are shown in Table 2. Prenatal perceived stress of the mother at T1 significantly predicted the change in the child’s emotional well-being (β= -0.018; *p* < 0.001), with higher PSS values at T1 being associated with a stronger decrease in children’s well-being during T5a compared to before the pandemic.

**Table 2.**
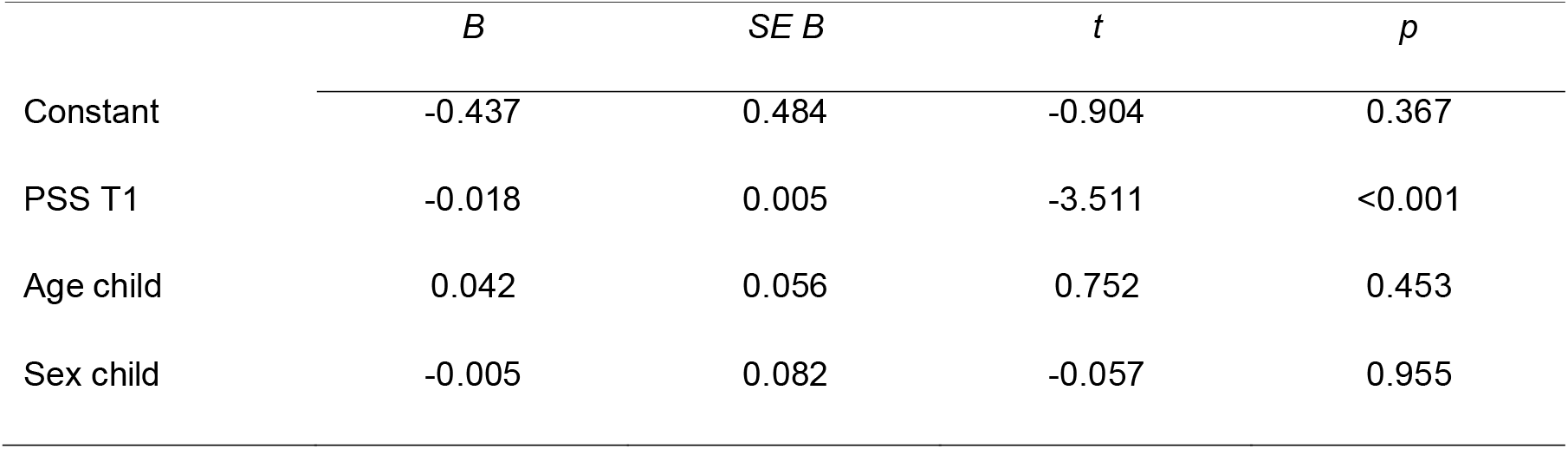
Parameter estimation to predict change in children’s emotional well-being

#### H3b

The model predicting the change in the mother’s emotional well-being during the pandemic with prenatal perceived stress at T1 was not significant (*F*(2, 191) = 1.229, *p* = 0.29, *R*^2^ = 0.013).

#### H3c

Furthermore predicting the change in the mother’s emotional well-being during the pandemic with perceived stress at T4 was not significant (*F*(2, 218) = 0.89, *p* = 0.41, *R*^*2*^ = 0.008)

### 3.5 H4: Association between HPA axis reactivity and the change in the child’s emotional well-being

The model predicting the change in the child’s emotional well-being with cortisol concentration in morning urine was not significant (*F*(3, 221) = 0.46; *p* = 0.71, *R* ^2^ = 0.006). The model predicting the change in the child’s emotional well-being with AUCi was not significant (*F*(2, 201) = 0.51; *p* = 0.60, *R*^2^ = 0.005).

### 3.6 H5: Association between polygenic risk for MDD, schizophrenia and loneliness and the change in the child’s emotional well-being

The change in the child’s emotional well-being during the pandemic could not be predicted by PRS for depression (*F*(8, 242) = 0.62; *p* = 0.76, *R*^2^ = 0.02), schizophrenia (*F*(8, 242) = 0.64; *p* = 0.74, *R*^2^ = 0.02) or loneliness (*F*(8, 242) = 0.65; *p* = 0.73, *R*^2^ = 0.02).

## Discussion

The present study investigated the well-being of children and adults during the COVID-19 pandemic, as well as its relation to prenatal perceived stress, early childhood HPA axis activity, and polygenic risk.

Consistent with our hypotheses (H1), the emotional well-being of children and mothers worsened during the pandemic compared to before the pandemic. The children’s and the mothers’ emotional well-being worsened at the beginning of the pandemic compared to pre-pandemic but the children’s emotional well-being improved during the second part of this study wave. Other studies have also shown that emotional well-being worsened in adults (Adams et al., 2021; Daly et al., 2020; Fancourt et al., 2021) and children (Achterberg et al., 2021; Bignardi et al., 2020; Ravens-Sieberer et al., 2020) at the beginning of the pandemic. There have been inconsistent findings considering emotional well-being over the course of the pandemic. Our results are in line with previous results showing improvement of emotional well-being among children (Achterberg et al., 2021; Skripkauskaite et al., 2020) over the course.

It is important to consider the circumstances, e.g., whether there was a lockdown and how strict restrictions were. Restrictions during the first assessment in June and July 2020, were already partly eased in Germany. During the second assessment beginning in November 2020, the second lockdown began, accompanied by more restrictions. These were comparable to those during the first assessment. On account of increasing coronavirus infections, a “hard lockdown” followed in December. Therefore, adults and children were affected by more restricting circumstances during the second assessment compared to the first. In addition, seasonal effects may have negatively affected emotional well-being, as the second assessment took place in winter, from November 2020 to February 2021 (Meesters and Gordijn, 2016). Another aspect could be that mothers already had additional difficulties with parenting (Spinelli et al., 2020) for a long period since the first lockdown. The combination of strict restrictions, depressive symptoms caused by seasonal effects and parenting difficulties could explain a worse emotional well-being at both assessed time points during the COVID pandemic.

Interestingly, children had a better emotional well-being in the second assessment compared to the first. This finding might be explained by adaptation to their environment and developing better coping mechanisms over the course of the pandemic. Also the social environment, e.g., families and schools, could have developed adapted routines, promoted resilience and had better strategies to support children compared to the beginning of the pandemic (Masten and Motti-Stefanidi, 2020). Future studies should evaluate which specific factors are helpful to strengthen children’s resilience.

A moderate association between the emotional well-being of the mother and the child during the pandemic (H2) was confirmed. This association has been shown by several studies (Russell et al., 2020; Spinelli et al., 2020) underlining that parent’s well-being is a mediator for children’s emotional and behavioral problems (Achterberg et al., 2021). Especially during stressful times like the COVID-19 pandemic, parents play an important role by influencing the mental health of their children (Russell et al., 2020). Previous findings have shown that prenatal stress has an impact on children’s development and health (Van den Bergh et al., 2020). Our findings suggest that prenatal maternal stress has an impact on children’s well-being several years later in life, particularly under stressful circumstances such as the COVID-19 pandemic (H3a). Recent studies have examined the association between prenatal stress and children’s mental health but starting assessment in the pregnancy during the pandemic (Buthmann et al., 2022; Duguay et al., 2022; Provenzi et al., 2021), therefore examining children much younger than the ones investigated in the present sample of 7-10 years old children. In order to examine how children’s development and mental health are affected by prenatal maternal health later in life, subsequent study waves should investigate this association in the present sample at a later stages.

There was no significant association between the perceived stress of the mother assessed in previous study waves and the change in the emotional well-being of the mother (H3b and c).

Results from another longitudinal study suggest that increases in depression and anxiety symptoms during the COVID-19 pandemic in mothers occurred universally, regardless of previous mental health history (Racine et al., 2021). Depending on current life circumstances, the amount of stress experienced, its perception and response to it can vary. For example during pregnancy (T1) and when children are at preschool age (T4) different stressors could have impacted the mother’s perceived stress. These stressors could be limited to the existing circumstances at the assessed time.

There was no significant association between the change in the emotional well-being of the child and the cortisol stress reactivity at 45 months (H4). In the same longitudinal birth cohort, prenatal stress was associated with a hyporegulation of the children’s HPA axis, as indicated by lower cortisol levels after a stress test (Send et al., 2019a) and lower cortisol and cortisone levels in the first morning urine of the then 45 month-old children (Send et al., 2019b). Various factors influencing the emotional well-being during the COVID-19 pandemic could lead to intraindividual changes, such as HPA axis reactivity. Interestingly, in other studies loneliness during the COVID-19 pandemic had been associated with higher levels of cortisol (Haucke et al., 2022; Jopling et al., 2021). Pandemic related circumstances may influence diurnal cortisol patterns. Including cortisol measurements in future assessments, allows us to explore the development of the HPA axis reactivity during childhood and adolescence.

PRSs did not significantly predict the change in the child’s emotional well-being during the pandemic (H5). It has to be taken into account that the application of PRSs is limited to the underlying GWAS and that to date even large GWASs in psychiatric genetics are still underpowered (Wray et al., 2014). Also, environmental factors such as isolation and school closures could have a larger effect on well-being than genetic risk factors.

The present study has several limitations. First, contrary to the earlier assessment waves, data acquisition during the pandemic only took place online or via mailed questionnaires. Second, the children did not answer the questions themselves. The main parent caregiver, in most cases the mother, answered the questions about the child. There is possible bias as answers about the emotional well-being of the child are influenced by the caregiver’s perspective and his/her own mental distress (De Los Reyes and Kazdin, 2005). In particular, the observed association between the mother’s and the child’s well-being could be partially driven by that bias. Third, there were only two assessments during the pandemic. It would be interesting to pursue the course of mental health linked to the prevailing circumstances. Fourth, the questionnaires assessing the three months prior to the pandemic were answered retrospectively, therefore that assessment is prone to recall biases. Fifth, our sample had limited diversity (mostly high educational background and high socio-economic status) and may not be representative of the general population. In line with this, it has to be noted that the levels of PSS assessed at T1 and T4 indicate that the mothers did not experience high levels of stress on average. Sixth, while we investigated a set of variables of interest, and adjusted our analysis for relevant technical covariates, and age and sex of the children, further variables might have influenced the observed results, such as actual COVID-19 infections, having lost the job or significant persons; or other measures on the child such as resilience, quality of attachment or cognitive aspects. Moreover, the PSS score of the mothers during the pandemic was not assessed and only previous assessed PSS measures could be included in the analysis. Additionally, as there was less data available from fathers than from mothers, the present study only included the mothers’ data. Future studies should additionally investigate the paternal perception of the pandemic.

The present part of the longitudinal birth cohort study assesses children’s emotional well-being during the COVID-19 pandemic and its’ association to stress linked predictors. The use of internationally established and validated screening instruments to assess mental health enables comparison with other studies. The study contributes to the emerging evidence that mental health of children and adults is affected by the COVID-19 pandemic. Furthermore, it indicates that the well-being of the children and their mothers was linked at each assessed time point. Moreover the results suggest that higher prenatal maternal stress is associated with a stronger decrease in children’s well-being at the age of 7-9 years. Due to the longitudinal study model, it is possible to set findings in relation to previous assessments of this cohort. While in the present study, associations with chosen genetic and endocrine predictors were not significant, differential results might be observed in children and adolescents of different age groups. Additionally, while not significant, these results add to the literature, where significant results might be overrepresented, e.g., due to publication bias. Further, it is of interest to investigate whether the experience of the pandemic has an impact on the health and development of the children in the future, especially in regard to their stress perception and endocrine stress regulation.

## Data Availability

All data produced in the present study are available upon reasonable request to the authors.

## Author contributions

Substantial contributions to the conception or design of the study, or the acquisition of data, or analysis and interpretation of data: T.N., L.Z, M.C., A.S.M.H., J.C.F., L.S., J.F., T.S.S., M.G., M.R., M.D., F.S, S.H.W.; Drafting the article or revising it critically for important intellectual content: T.N., L.Z., J.C.F., S.H.W., F.S; All authors approved the final version of the manuscript.

## Acknowledgements

We thank all parents and children for taking part in this study and our student employees and interns for their support with data acquisition and data entry

